# Strength-Duration Curve Outcomes, Profiles and Evidence in Spinal Cord Injury: Protocol for a Scoping Review (SCOPE-SCI Study)

**DOI:** 10.1101/2025.09.08.25335310

**Authors:** Ishmeet Singh, Ankush Gera, Twinkle Luthra, Ishant Arora

**Affiliations:** Cambridgeshire and Peterborough NHS Foundation Trust, Cambridge, United Kingdom; Sri Balaji Action Medical Institute, New Delhi, India; Jamia Millia Islamia Institute, New Delhi, India

## Abstract

**Introduction:** Neuromuscular and functional electrical stimulation (NMES/FES) are widely used in spinal cord injury (SCI) rehabilitation to preserve muscle health, manage spasticity, and support functional recovery. However, stimulation protocols typically use fixed parameters with limited individualisation. The strength–duration (SD) curve, reflecting the relationship between pulse duration and stimulus intensity required for muscle contraction, captures excitability changes due to denervation, reinnervation, and training. While historically used in neurophysiology, its translational role in guiding NMES/FES programming in SCI rehabilitation remains unclear. This scoping review aims to map current evidence on SD-curve measurement, outcomes, and clinical applications in SCI.

**Methods and analysis:** This protocol follows PRISMA-ScR guidelines. We will search MEDLINE, EMBASE, CINAHL, and grey literature sources from inception to present. Eligible studies will include adults with traumatic or non-traumatic SCI, reporting SD curves or related excitability measures (rheobase, chronaxie, accommodation index). Data extraction will capture study characteristics, methods of SD-curve measurement, rheobase/chronaxie values, clinical applications (diagnostic, prognostic, therapeutic), and outcomes of SD-guided NMES/FES interventions. Results will be presented descriptively and thematically, with tables and figures mapping measurement approaches, reported values, and translational applications. Methodological and reporting gaps will be highlighted.

**Ethics and dissemination:** Ethical approval is not required. Findings will be disseminated through peer-reviewed publication, international conferences, and to inform future pilot studies on SD-curve–guided rehabilitation in SCI.

## Background and Rationale

Spinal cord injury (SCI) is a complex neurological condition leading to profound motor, sensory, and autonomic impairments with long-term functional and psychosocial consequences. Despite advances in acute management and rehabilitation, restoration of motor function remains a major challenge, particularly in chronic stages of injury where muscle atrophy, spasticity, and altered excitability complicate recovery pathways [1,2]. Rehabilitation strategies increasingly rely on neuromodulation and neurostimulation approaches such as neuromuscular electrical stimulation (NMES), functional electrical stimulation (FES), and epidural or cortical stimulation to maintain muscle health, enhance neuroplasticity, and promote functional recovery [3–5]. Optimizing stimulation parameters is critical to maximize efficacy, reduce patient discomfort, and ensure safety during long-term rehabilitation.

The strength–duration (SD) curve, a fundamental electrophysiological tool, characterizes the excitability of muscle and nerve tissue by quantifying the relationship between stimulus intensity and pulse duration. Derived indices such as rheobase and chronaxie provide insight into motor unit recruitment, denervation status, and changes in neuromuscular excitability [6,7]. Historically, SD curves have been employed to differentiate between upper motor neuron (UMN) and lower motor neuron (LMN) lesions and to guide electrical stimulation protocols in peripheral nerve injuries [8]. In neurological rehabilitation, they offer a potential framework for tailoring stimulation parameters, thereby improving treatment precision.

Emerging evidence suggests that SD curve profiling may hold diagnostic, prognostic, and therapeutic relevance across several neurological conditions. In stroke, SD curve alterations have been linked with motor recovery potential and response to FES interventions [9]. In multiple sclerosis and traumatic brain injury, changes in excitability parameters have been explored as indicators of disease-related neuromuscular dysfunction [10]. In SCI, however, application of SD curves remains limited, fragmented, and inconsistently reported. While some studies have documented altered rheobase and chronaxie values in paralyzed muscles, systematic application of SD curve findings to guide NMES or FES dosing remains underdeveloped [11–13].

Several methodological gaps hinder translation of SD curves into routine SCI rehabilitation. Variability in measurement techniques—such as differences in electrode size, waveform, pulse duration, and threshold definitions—complicates comparisons across studies [14]. Safety considerations, particularly for long-pulse or galvanic stimulation, are inconsistently addressed, raising concerns regarding skin irritation or tissue damage [15]. Moreover, the majority of studies are cross-sectional, with limited exploration of longitudinal SD curve monitoring to track recovery or inform adaptive stimulation protocols. The lack of integration of SD curve–guided NMES/FES parameterization into clinical trials further limits evidence on its contribution to functional recovery, muscle excitability, and patient tolerance.

Given these gaps, there is a pressing need to systematically map and synthesize evidence on the application of SD curves in SCI and related central nervous system (CNS) disorders. Such synthesis will clarify existing methods, document reported rheobase/chronaxie ranges, and examine the extent to which SD curves have informed NMES/FES programming and rehabilitation outcomes. Importantly, identifying methodological limitations and safety concerns will guide future development of adaptive, closed-loop, and personalized stimulation strategies in SCI rehabilitation.

### Review Objectives

The following are the objectives of the review-

1. To systematically review the methods of SD-curve measurement in individuals with SCI and related CNS disorders, including variations in pulse width, electrode size, waveform, threshold definitions, and reporting practices, along with safety limits and monitoring protocols for long-pulse/galvanic stimulation.
2. To synthesise reported rheobase and chronaxie values across upper motor neuron (UMN), lower motor neuron (LMN), and mixed lesion presentations, and to explore how these electrophysiological parameters vary with injury level, severity, chronicity, and therapeutic interventions.
3. To examine the clinical applications of SD curves in SCI, including their use in diagnostic (assessment of denervation and excitability status), prognostic (prediction of recovery potential), and therapeutic contexts (guiding NMES/FES, CES, and other neuromodulation approaches).
4. To evaluate how SD-curve findings have been applied to optimize stimulation parameters (pulse duration, frequency, intensity, duty cycle) for NMES/FES and related interventions, and the outcomes associated with such applications, including muscle excitability, contractile responses, functional recovery, and patient tolerance.
5. To identify methodological limitations and reporting gaps, and to highlight opportunities for innovation-particularly the longitudinal use of SD curves to track recovery, and the development of adaptive or closed-loop SD-curve–based strategies for personalized rehabilitation in SCI.

### Review Questions

1. How have SD curves been measured in individuals with SCI (methods, safety, protocols)?
2. What rheobase and chronaxie values are reported in SCI populations, and how do they vary across lesion types, levels, chronicity, and interventions?
3. In what contexts (diagnostic, prognostic, therapeutic) have SD curves been applied in SCI?
4. How have SD-curve findings informed NMES/FES settings, and what outcomes have been reported?
5. What methodological and reporting gaps exist, and what directions are indicated for future research and clinical translation?

### Eligibility Criteria (PCC Framework)

#### Population

Adults (≥18 years) with traumatic or non-traumatic spinal cord injury (SCI), at any neurological level of injury or severity (AIS A–D). Studies including mixed CNS populations (e.g., stroke, MS) will be eligible only if SCI subgroup data are extractable.

#### Concept

Studies assessing SD curves or related excitability measures (rheobase, chronaxie, accommodation index, excitability time constants) for diagnostic, prognostic, or therapeutic purposes.

#### Context

Rehabilitation, clinical, or experimental settings, across all injury stages (acute, subacute, chronic).

#### Inclusion Criteria

- Empirical studies of any design: Randomized controlled trials, quasi-experimental, cohort, cross-sectional, case–control, case series, and case reports.
- Grey literature where methodology is described (e.g., thesis, dissertations, technical reports, conference proceedings).
- All years of publication.
- English-language studies (translation not feasible within scope).

#### Exclusion Criteria

- Animal-only studies.
- Non-SCI populations without separable SCI data.
- Purely theoretical, modeling, or simulation papers without human SCI data.
- Studies limited to **electromyography or neurophysiological measures** unrelated to SD-curve derivation.
- Reviews, editorials, or commentaries (though reference lists will be screened for eligible studies).

### Search Strategy

#### Population (SCI)

- “spinal cord injur*” OR SCI OR paraplegi* OR tetraplegi* OR quadriplegi* OR myelopath*
- [MeSH/Emtree terms]: *Spinal Cord Injuries*

#### Concept (SD-curve / excitability)

- “strength duration” OR “SD curve*” OR “excitability curve*” OR rheobase OR chronaxie OR “stimulus duration” OR “stimulus response” OR “accommodation index” OR “nerve excitability”
- [MeSH/Emtree terms]: *Electrophysiology* OR *Electric Stimulation* OR *Action Potentials*

#### Context (NMES/FES/rehab)

- “neuromuscular electrical stimulation” OR NMES OR FES OR CES OR “functional electrical stimulation” OR “electrical muscle stimulation” OR “galvanic stimulation” OR “long pulse stimulation” OR electrotherap* OR rehabilitat*

Searches will be supplemented by reference screening, citation tracking, and grey literature review. A full Boolean search string will be provided for MEDLINE.

### Study Selection

Titles and abstracts will be screened independently by two reviewers. Full texts will be retrieved for potentially relevant studies and assessed in duplicate. Conflicts will be resolved by consensus or a third reviewer. The selection process will be documented using the PRISMA 2020 flow diagram, with reasons for exclusion recorded. Screening will be piloted on a random sample before formal selection. An appropriate screening tool will be used to support the process.

### Data Charting (Extraction)

A standardized, piloted data-charting form will be used. The following domains will be extracted from eligible studies:

- **Bibliographic details:** Author(s), year, country, journal/source, study design.
- **Population characteristics:** Sample size, participant demographics (age, sex where available), SCI etiology (traumatic/non-traumatic), injury level, severity (AIS), and chronicity.
- **SD-curve methodology:** Muscles tested, stimulation device/equipment, electrode type/size/placement, waveform, pulse widths tested, threshold definition, measurement protocol, rheobase and chronaxie values, reporting of excitability/trophic stimulation.
- **Context and application:** Purpose of SD-curve use (diagnostic, prognostic, therapeutic, or monitoring), and study setting (rehabilitation, clinical, experimental, or epidural stimulation).
- **Stimulation parameters (FES/NMES/CES):** Frequency, amplitude, pulse duration, duty cycle, electrode configuration, parameter adjustments guided by SD-curve findings.

- **Outcomes assessed**
  ∘ *Physiological:* muscle excitability, EMG activity, contractile responses, trophic changes.
  ∘ *Clinical/functional:* strength, motor recovery, functional performance.
  ∘ *Patient-reported/safety:* tolerance, comfort, adverse events, monitoring protocols.

- **Key findings and implications:** Main results, relevance for rehabilitation and stimulation practice, methodological limitations, and identified research gaps.

### Data Synthesis

The extracted data will be synthesised using descriptive and thematic analysis. Findings will be organized into tabular and narrative summaries to allow systematic mapping of the evidence.

The synthesis will include:

- **Measurement methods:** stimulation devices, electrode characteristics, pulse parameters, muscles studied, and threshold definitions.
- **Reported values:** ranges of rheobase and chronaxie, stratified by lesion type (UMN, LMN, or mixed), injury stage (acute, subacute, chronic), and therapeutic intervention exposure.
- **Applications:** diagnostic (denervation status), prognostic (prediction of recovery), and therapeutic uses (guiding NMES/FES/CES protocols).
- **Stimulation protocols informed by SD-curves:** adjustments in frequency, amplitude, and pulse duration, and their relation to physiological and clinical outcomes.
- **Outcomes:** muscle excitability, EMG responses, strength, functional recovery, patient tolerance, and adverse events.
- **Safety and monitoring:** reported safety limits, adverse effects, and monitoring protocols for long-pulse or galvanic stimulation.
- **Evidence gaps:** absence of longitudinal studies, inconsistent reporting of functional outcomes, methodological heterogeneity, and lack of adaptive or closed-loop applications.

Narrative mapping will emphasize consistency and variability across studies, and identify opportunities for methodological refinement and future research.

## Data Availability

All data produced are available online at

https://doi.org/10.17605/OSF.IO/DU9GN

## Funding

No funding has been secured for this review.

## Ethics and Dissemination

As a review of published literature, no ethical approval is required. Findings will be disseminated via peer-reviewed publication, international conference presentations (ISCoS, ISSICON etc.), and integration into a subsequent longitudinal pilot study design.

## Review Registration

This protocol will be prospectively registered on the Open Science Framework (OSF) and/or as a preprint on medRxiv. The review protocol will also be submitted for consideration in a peer reviewed journal.

The review will be conducted in line with the PRISMA-ScR guidelines (Tricco et al., 2018).

## Appendices

### Appendices

#### 1. Boolean search string for MEDLINE (via PubMed)

(spinal cord injur*[Title/Abstract] OR SCI[Title/Abstract] OR

paraplegi*[Title/Abstract] OR tetraplegi*[Title/Abstract]

OR quadriplegi*[Title/Abstract] OR myelopath*[Title/Abstract]

OR “Spinal Cord Injuries”[MeSH])

AND

(“strength duration”[Title/Abstract] OR “SD curve*”[Title/Abstract] OR “excitability curve*”[Title/Abstract]

OR rheobase[Title/Abstract] OR chronaxie[Title/Abstract] OR “stimulus duration”[Title/Abstract]

OR “stimulus response”[Title/Abstract] OR “accommodation index”[Title/Abstract]

OR “Electrophysiology”[MeSH] OR “Electric Stimulation”[MeSH] OR “Action Potentials”[MeSH])

AND

(“neuromuscular electrical stimulation”[Title/Abstract] OR NMES[Title/Abstract]

OR “functional electrical stimulation”[Title/Abstract] OR FES[Title/Abstract]

OR CES[Title/Abstract] OR “electrical muscle stimulation”[Title/Abstract]

OR “galvanic stimulation”[Title/Abstract] OR “long pulse stimulation”[Title/Abstract]

OR electrotherap*[Title/Abstract] OR rehabilitat*[Title/Abstract])

2. EMBASE

(‘spinal cord injury’/exp OR ‘spinal cord injur*’:ti,ab OR SCI:ti,ab OR paraplegi*:ti,ab OR tetraplegi*:ti,ab OR quadriplegi*:ti,ab OR myelopath*:ti,ab)

AND

(‘strength duration’:ti,ab OR ‘SD curve*’:ti,ab OR ‘excitability curve*’:ti,ab OR rheobase:ti,ab OR chronaxie:ti,ab

OR ‘stimulus duration’:ti,ab OR ‘stimulus response’:ti,ab OR ‘accommodation index’:ti,ab

OR ‘electrophysiology’/exp OR ‘electric stimulation’/exp OR ‘action potentials’/exp)

AND

(‘neuromuscular electrical stimulation’:ti,ab OR NMES:ti,ab OR ‘functional electrical stimulation’:ti,ab OR FES:ti,ab

OR CES:ti,ab OR ‘electrical muscle stimulation’:ti,ab OR ‘galvanic stimulation’:ti,ab OR ‘long pulse stimulation’:ti,ab

OR electrotherap*:ti,ab OR rehabilitat*:ti,ab)

#### 3. CINAHL (EBSCOhost)

((MH “Spinal Cord Injuries+”) OR TI “spinal cord injur*” OR AB “spinal cord injur*” OR TI SCI OR AB SCI

OR TI paraplegi* OR AB paraplegi* OR TI tetraplegi* OR AB tetraplegi* OR TI quadriplegi* OR AB quadriplegi* OR TI myelopath* OR AB myelopath*)

AND

(TI “strength duration” OR AB “strength duration” OR TI “SD curve*” OR AB “SD curve*” OR TI “excitability curve*” OR AB “excitability curve*”

OR TI rheobase OR AB rheobase OR TI chronaxie OR AB chronaxie OR TI “stimulus duration” OR AB “stimulus duration”

OR TI “stimulus response” OR AB “stimulus response” OR TI “accommodation index” OR AB “accommodation index”

OR (MH “Electrophysiology”) OR (MH “Electric Stimulation”) OR (MH “Action Potentials”))

AND

(TI “neuromuscular electrical stimulation” OR AB “neuromuscular electrical stimulation” OR TI NMES OR AB NMES

OR TI “functional electrical stimulation” OR AB “functional electrical stimulation” OR TI FES OR AB FES

OR TI CES OR AB CES OR TI “electrical muscle stimulation” OR AB “electrical muscle stimulation”

OR TI “galvanic stimulation” OR AB “galvanic stimulation” OR TI “long pulse stimulation” OR AB “long pulse stimulation”

OR TI electrotherap* OR AB electrotherap* OR TI rehabilitat* OR AB rehabilitat*)

## Notes

### Competing Interest Statement

The authors have declared no competing interest.

### Clinical Protocols

https://doi.org/10.17605/OSF.IO/DU9GN

### Funding Statement

This study did not receive any funding

## References

1. Ahuja CS, Wilson JR, Nori S, Kotter MR, Druschel C, Curt A, et al. Traumatic spinal cord injury. Nat Rev Dis Primers. 2017;3:17018.

2. Kirshblum SC, Botticello A, Lammertse D, Marino R, Chiodo A, Jha A, et al. Characterizing natural recovery after traumatic spinal cord injury. J Neurotrauma. 2021;38(9):1267–74.

3. Wagner FB, Mignardot JB, Le Goff-Mignardot CG, Demesmaeker R, Komi S, Capogrosso M, et al. Targeted neurotechnology restores walking in humans with spinal cord injury. Nature. 2018;563(7729):65–71.

4. Ho CH, Triolo RJ, Elias AL, Kilgore KL, DiMarco AF, Bogie K, et al. Functional electrical stimulation and spinal cord injury. Phys Med Rehabil Clin N Am. 2014;25(3):631–54.

5. Gad P, Gerasimenko Y, Zdunowski S, Turner A, Sayenko D, Lu DC, et al. Iron neuroprosthesis for recovery after spinal cord injury. J Neurotrauma. 2015;32(9):701–9.

6. Irnich W. The chronaxie time and its practical importance. Pacing Clin Electrophysiol. 1980;3(2):292–301.

7. Bostock H, Cikurel K, Burke D. Threshold tracking techniques in the study of human peripheral nerve. Muscle Nerve. 1998;21(2):137–58.

8. Kralj A, Bajd T, Turk R. Electrical stimulation providing functional use of paraplegic patient muscles. Med Prog Technol. 1980;7(1):3–9.

9. Hara Y, Obayashi S, Tsujiuchi K, Muraoka Y. The effects of electromyography-controlled functional electrical stimulation on upper limb function in stroke patients: a randomized controlled trial. Clin Rehabil. 2013;27(6):579–87.

10. Humm AM, Beer S, Kool J, Magistris MR, Kesselring J, Rösler KM. Quantification of axonal loss in multiple sclerosis with motor evoked potentials. J Neurol. 2004;251(11):1385–94.

11. Petrofsky JS. Electrical stimulation: neurophysiological basis and application. Basic Appl Myol. 2004;14(4):205–13.

12. Mayr W, Bijak M, Rafolt D, Sauermann S, Unger E, Lanmüller H. Basic design and construction of functional electrical stimulation (FES) devices for therapy and research. Artif Organs. 2001;25(3):206–9.

13. Gobbo M, Maffiuletti NA, Orizio C, Minetto MA. Muscle motor point identification is essential for optimizing neuromuscular electrical stimulation use. J Neuroeng Rehabil. 2014;11:17.

14. de Freitas GR, Marcolino AM, Silva TM, Binda AC, Nogueira-Neto GN, Polacow MLO. Strength-duration curves in denervated muscles: methodological aspects and clinical implications. Rev Bras Fisioter. 2010;14(6):503–9.

15. Peckham PH, Knutson JS. Functional electrical stimulation for neuromuscular applications. Annu Rev Biomed Eng. 2005;7:327–60.

